# The Moderating Role of Genetic and Environmental Risk Factors for Schizophrenia on the Relationship between Autistic Traits and Psychosis Expression in the General Population

**DOI:** 10.1101/2024.11.21.24317723

**Authors:** Melike Karacam Dogan, Thanavadee Prachason, Bochao Lin, Lotta-Katrin Pries, Angelo Arias-Magnasco, Riccardo Bortoletto, Claudia Menne-Lothmann, Jeroen Decoster, Ruud van Winkel, Dina Collip, Philippe Delespaul, Marc De Hert, Catherine Derom, Evert Thiery, Nele Jacobs, Jim van Os, Bart Rutten, Natascia Brondino, Marco Colizzi, Jurjen Luykx, Laura Fusar-Poli, Sinan Guloksuz

**Affiliations:** Department of Psychiatry, Karadeniz Eregli State Hospital, Zonguldak, Turkey; Department of Psychiatry, Faculty of Medicine, Ramathibodi Hospital, Mahidol University, Bangkok, Thailand; Department of Psychiatry and Neuropsychology, School for Mental Health and Neuroscience, Maastricht University Medical Center, Maastricht, the Netherlands; Unit of Psychiatry, Department of Medicine (DMED), University of Udine, Udine, Italy; University Psychiatric Centre KU Leuven, KU Leuven, Belgium; Centre of Human Genetics, University Hospitals Leuven, Belgium; Department of Neurology, Ghent University Hospital, Ghent, Belgium; Faculty of Psychology and Educational Sciences, Open University of the Netherlands; UMC Utrecht, Division Neuroscience, Utrecht, the Netherlands; Department of Psychosis Studies, Institute of Psychiatry, King’s Health Partners, King’s College London, London, UK; Department of Brain and Behavioral Sciences, University of Pavia, Pavia, Italy; Amsterdam University Medical Centres, Department of Psychiatry; Department of Psychiatry, Yale School of Medicine, New Haven, CT, USA

**Keywords:** autistic traits, psychotic experiences, genetic risk factors, environmental risk factors, exposome, childhood trauma, polygenic risk scores

## Abstract

**Background and Hypothesis:** Psychosis-related environmental risks are common in autism, with identified genetic connections to psychosis. Therefore, we hypothesized these risks may moderate the relationship between autistic traits (ATs) and psychotic experiences (PEs).

**Study Design:** First-wave data from 792 twins and siblings in the TwinssCan Project were analysed (n=792). PEs and ATs were assessed using the Community Assessment of Psychic Experiences and the Autism-Spectrum Quotient. Polygenic risk scores for schizophrenia and psychosis-associated environmental factors (childhood trauma (CT), bullying, negative life events, obstetric complications, cannabis use, winter birth, and hearing impairment) were tested for independent and interactive effects with ATs using separate multilevel linear regression models.

**Study Results:** ATs, all five CT subtypes, bullying, and negative life events were significantly associated with PEs (all P < 0.004). Emotional abuse (B: 0.08, 95% CI:0.05 to 0.11, P < 0.001), physical abuse (B: 0.11, 95% CI: 0.05 to 0.18, P = 0.001), sexual abuse (B: 0.09, 95% CI: 0.03 to 0.15, P = 0.002), and physical neglect (B: 0.06, 95% CI: 0.03 to 0.10, P = 0.001) significantly amplified the positive relationship between ATs and PEs, whereas emotional neglect (B: 0.04, 95% CI: 0.01 to 0.07, P = 0.007) and negative life events (B: 0.007, 95% CI: 0.0005 to 0.014, P = 0.04) only showed a trend of interactions. No significant main or interacting effects of genetic and other risk factors were found.

**Conclusions:** These findings imply that CT might be a potential preventive target for psychosis expression in people with high ATs.

## Introduction

Autism Spectrum Disorder (ASD) is a complex neurodevelopmental condition characterized by persistent deficits in social communication and social interaction along with restricted, repetitive patterns of behaviors, interests, and/or activities^1^. The prevalence of ASD has recently been estimated to be 1 in 36 children in the United States^2^. Psychiatric comorbidity is common in ASD. The prevalence of comorbid conditions has been estimated to be 28% for attention-deficit hyperactivity disorder (ADHD), 20% for anxiety disorders, 11% for depressive disorders, and 9% for schizophrenia spectrum disorders^3^. More recent meta-analyses of studies in autistic people consistently reported a pooled prevalence of 9.4% for clinical psychosis^4^ and 24% for psychotic experiences (PEs)^5^, highlighting the significance of clinical and subclinical psychosis among commonly co-occurring conditions in ASD.

In the past two decades, autism has increasingly been recognized as a true spectrum with varying degrees of traits existing on a continuum in the general population. According to such a concept, autistic traits (ATs), such as social communication difficulties and repetitive behaviors, could also be present among individuals who do not meet the formal diagnostic criteria for ASD. Similar to clinical ASD, ATs have been shown to be associated with an increased vulnerability to multiple psychopathologies, including subclinical psychosis expression^6–9^. A recent meta-analysis has identified multiple demographic and clinical characteristics, such as older age, male gender, lower intellectual quotient, and less stereotyped interests/behaviors, as risk factors for developing clinical psychosis among autistic individuals^4^. Nevertheless, evidence on risk factors for subclinical psychosis among people with subthreshold autism is lacking.

Considering that genetic and environmental risk factors for schizophrenia are shared across the psychosis continuum^10^ and throughout a neurodevelopmental trajectory also enclosing other conditions emerging earlier in life^11^, it is likely that the risk factors may also influence psychosis expression in the context of the autism spectrum. In the present study, we, therefore, aimed to examine the moderating role of polygenic risk and well-established environmental risk factors for schizophrenia (i.e., childhood trauma, bullying, negative life events, obstetric complications, cannabis use, winter birth, and hearing impairment) on the relationship between ATs and PEs in the general population. We hypothesized that the risk factors for schizophrenia would amplify the strength of the association between ATs and PEs.

## Methods

### Participants

Analyses were conducted on the dataset obtained from the first wave of the TwinssCan Project. The details of participant enrollment and data collection have been previously described elsewhere^12^. Briefly, the TwinssCan cohort comprises twins, non-twin siblings, and parents recruited from the East Flanders Prospective Twin Survey (EFPTS), a prospective population-based registry of multiple births from East Flanders, Belgium^13^. The twins were between 15 and 18 years old upon enrollment, while their non-twin siblings were from 15 to 35 years old. Written informed consent was obtained from all participants. The exclusion criteria were: (1) lack of parental or caregiver consent for those under 18 years old and (2) having a pervasive mental disorder. The study was approved by the local ethics committee (Commissie Medische Ethiek van de Universitaire Ziekenhuizen KU Leuven, Nr. B32220107766). In this study the data of 821 twins and siblings were included. However, 29 participants were excluded due to missing data, leaving 792 participants for further analysis.

### Measures

#### Psychotic Experiences

PEs were assessed with the Community Assessment of Psychic Experiences (CAPE), a widely used 42-item self-report questionnaire consisting of three subscales (i.e., positive, negative, and depressive)^14^. The CAPE has demonstrated stability, reliability, and validity in evaluating PEs in the general population^15^. Four-point Likert scales were used to measure the frequency and distress levels of the symptoms. The average scores from these scales give total and subscale scores (positive, negative, depressive). In our analysis, we specifically used the frequency scores as an indicator of PEs.

#### Autistic Traits

The Autism-Spectrum Quotient (AQ), a 50-item self-report questionnaire, was used to assess ATs^16^. It is widely used to measure ATs in the general population and has good psychometric properties through comprehensive research^16,17^. The scale consists of 5 subdomains, each with 10 items: imagination, social skills, communication, attention to detail, and attention shifting.

For items in line with ATs, “definitely agree” and “slightly agree” responses were scored 1, and “definitely disagree” and “slightly disagree” responses were scored 0^16^. For the other items, “definitely disagree” and “slightly disagree” were reversely scored 1; otherwise, 0. The total score ranges from 0 to 50 with higher AQ scores indicating higher ATs^16^.

#### Childhood Trauma

Childhood trauma (CT) was assessed using the Childhood Trauma Questionnaire (CTQ)^18^, a 28-item self-report instrument rated on a 5-point Likert scale to measure five subtypes of childhood adversity: emotional, physical, and sexual abuse, as well as emotional and physical neglect. In the manual of CTQ, three severity cut-off scores (low, moderate, and severe) were suggested for each CTQ subscale^19^. Aligning with previous research^20,21^, we binarized the variable into the presence or absence of each CT subtype by employing the low severity cut- off threshold based on the manual of the CTQ (≥ 9 for emotional abuse, ≥ 8 for physical abuse, ≥ 6 for sexual abuse, ≥ 10 for emotional neglect, and ≥ 8 for physical neglect). These cut-off scores were used to maintain consistency and increase comparability across studies.

#### Bullying

The Retrospective Bullying Questionnaire (RBQ) was used to assess the experiences of bullying before the age of 17^22^. It is a 44-item self-rated questionnaire examining different aspects of bullying, including verbal, physical, and relational aggression, experienced across settings like school, neighborhood, and online platforms. The severity of different kinds of bullying was rated on a 5-point Likert scale (1=not being bullied, 2=not serious at all, 3=just a little, 4=quite serious, 5=extremely serious). Aligning with prior research, bullying victimization was considered present if they had encountered any type of bullying (cut-off score≥ 2)^23^.

#### Negative life events

Negative life events were assessed with the Life Events Questionnaire (LEQ), a 61-item self-report of major life events such as job loss, illness, death of a loved one, and financial changes^24^. Participants indicated if these incidents had ever happened to them. The total score was calculated by summing the number of negative life events experienced over a lifetime.

#### Obstetric complications

Adverse events during pregnancy, labor, and postpartum period were measured with the McNeil-Sjöström Obstetric Complications (OC) Scale (MSS)^25^. The severity of OC was scored on a 6-point scale (1 = not harmful or relevant to 6 = very great harm or deviation in offspring). Aligning with prior research^26^, a dichotomous OC variable was generated using an MSS cut- off score of 5 or more to indicate severe OC. In other words, severe OC was defined as 1) birth weight lower than 2 kilograms, 2) birth weight more than 20% lower than that of the sibling, 3) version extraction delivery mode, 4) face or forehead presentation of the fetus during delivery, or 5) umbilical cord complication.

#### Cannabis use

Lifetime cannabis use was assessed by the L section of Composite International Diagnostic Interview (CIDI)^27^. A dichotomous variable was generated to indicate cannabis use over a lifetime.

#### Winter birth

The high-risk birth period was defined as the winter solstice (December-March) based on previous studies investigating the link between birth season and schizophrenia spectrum disorders in the Northern Hemisphere^28^. A dichotomous variable was generated to indicate winter birth.

#### Hearing impairment

Hearing impairment was evaluated based on the participants’ self-reported current hearing status.

#### Polygenic risk score for schizophrenia (PRS-SZ)

##### Genotyping

As previously reported^10^, genotypes of the twins and their siblings were generated on two platforms: the Infinium CoreExome-24 and Infinium PsychArray-24 kits. Quality control (QC) procedures were performed using PLINK v1.9 in both datasets separately^29^. Single-nucleotide polymorphisms (SNPs) and participants with call rates below 95% and 98%, respectively, were removed. A strict SNP QC was conducted only for subsequent sample QC steps. This involved a minor allele frequency (MAF) threshold >10% and a Hardy–Weinberg equilibrium (HWE) P-value >10^−5^, followed by linkage disequilibrium (LD)-based SNP pruning (R ^2^<0.5). This resulted in ∼58K SNPs to assess sex errors (n=8), heterozygosity [F < mean - 5× the standard deviation (SD), n = 3], homozygosity (F > mean + 5× SD), and relatedness by pairwise identity by descent (IBD) values (monozygotic:*p̂* < *0*.*9*, dizygotic and full siblings: *p̂* > *0*.*65* or *p̂* < *0*.*35*, n = 5). The ancestry-informed principal component (PC) analyses were conducted using EIGENSTRAT^30^. The ethnic outliers of which the first 4 PCs diverged >10× SD from Utah residents with Northern and Western European ancestry from the Centre d’Etude du Polymorphisme Humain (CEPH) collection (Central EUrope, CEU) and Toscani in Italia (TSI) samples (n=5), and >3× SD of the TwinssCan samples (n=7) were excluded (see also supplementary figure 1). After removing these subjects, a regular SNP QC was performed (SNP call rate >98%, HWE P > 1e-06, MAF > 1%, and strand ambiguous SNPs and duplicate SNPs were removed).

The two QCed datasets were imputed on the Michigan server^31^, using the Haplotype Reference Consortium r1.1 2016 reference panel with European samples after phasing with Eagle v2.3. Post-imputation QC involved removing SNPs with imputation quality (*R* ^2^) < 0.8, with an MAF < 0.01, SNPs that had a discordant MAF compared to the reference panel (MAF difference with HRC reference > 0.15), as well as strand ambiguous AT/CG SNPs and multi-allelic SNPs. The two chips were merged, and an additional check for MAF > 0.01, HWE P > 1e-06 was executed, which resulted in 3,407,392 SNPs for 688 individuals. The general imputation quality is shown in Supplementary Figure 1.

##### PRS calculation

Twelve PRSs were calculated based on the most recent genome-wide association study (GWAS) for schizophrenia^32^. To ensure the ethnicity match between training and target datasets, we selected the summary statistics from the European population. PRScs-auto was used to infer PRS generated using posterior SNP effect sizes, by placing a continuous shrinkage (cs) prior on SNP weights reported in the summary statistics and combined with an external linkage disequilibrium reference panel, such as the 1000 Genomes Project European Sample (https://github.com/getian107/PRScs). For this process, we used the PRS-CS “auto” function, which employs a fully Bayesian approach. This approach automatically learns the global shrinkage parameter (ϕ) from the available data. Unlike other methods, it does not prune SNPs based on a specific p-value threshold or perform clumping for independent SNPs. Instead, it assumes a general distribution of effect sizes across the genome and considers LD between SNPs using an external LD reference panel (in our case, the 1000 Genomes Phase 3 European samples panel). To compute posterior effect sizes, the default settings of PRS-cs-auto were used. After the calculation of posterior effect sizes, PRSs were calculated using ‘--score’ function and the SUM modifier in PLINK 1.9. After quality control, 3,366,081 variants were used in the PRS calculation.

Besides PRScs-auto, we also calculated PRSice with the clumping and threshold methods using PRSice2 (Choi et al., 2020), for sensitivity analysis to verify the results. To calculate all PRSice, the beta-values, effective allele, and P-values were extracted from all summary statistics. Insertions and deletions, ambiguous SNPs, SNPs with a MAF<0.01 and/or imputation quality R^2^<0.9, as well as SNPs located in complex-LD regions and long-range LD regions^33^ were excluded from TwinssCan dataset (see Supplementary table 1). Overlapping SNPs between GWAS summary statistics (training dataset), 1000 genomes (reference), and our TwinssCan dataset (target) were selected. These SNPs were clumped in two rounds using PLINK’s clump function (round 1: --clump-kb 250 --clump-r2 0.5; round 2: --clump-kb 5000 --clump-r2 0.2). The numbers of alleles for PRS calculation are listed in Supplementary Table 2. Odds ratios for autosomal SNPs reported in the neuroticism summary statistics were log-converted into beta values. PRS was calculated using PRSice2 (Choi et al., 2020) at the following P-value thresholds: 5 x 10^-6^, 5 x 10^-5^, 5 x 10^-4^, 5 x 10^-3^, 0.05, 0.1, 0.2, 0.3, 0.4, 0.5 and 1.

The P-threshold of < 0.05 was used; since this threshold accounted for the most of the variation in the phenotype according to the analysis conducted by the Psychiatric Genomics Consortium.

### Statistical analysis

All analyses were performed using Stata 16^34^. Participants were clustered within twin pairs, prompting the use of multilevel analyses. Firstly, to examine the main association between risk factors with PEs, multilevel linear regression analyses were performed using total CAPE as the dependent variable and AQ scores, PRS-SZ, CT subtypes, bullying, negative life events, obstetric complications, cannabis use, or winter birth as an independent variable tested in separate models. To examine whether the psychosis-associated genetic and environmental risk factors moderate the association between ATs and symptoms, a full interaction of total AQ and genetic or each environmental risk factor on total CAPE was tested using multilevel linear regression analyses. To facilitate the interpretation of the coefficients, AQ scores, negative life events, and PRS-SZ were standardized to have a mean of 0 and standard deviation (SD) of 1. All models were adjusted for age and gender. The models with PRS-SZ as an independent variable were additionally adjusted for the first two genomic PCs in accordance with previous research in this sample^35^. The statistical significance threshold was set at Bonferroni-corrected P< 0.004. As exploratory analyses, similar multilevel linear regression analyses were performed using CAPE subscales (i.e., positive, negative, and depressive subscales) as dependent variables.

## Results

### Sample characteristics

A total of 792 participants, including 274 monozygotic twins, 475 dizygotic twins, and 43 siblings, were included in the current analyses. Participant characteristics are displayed in Table 1.

**Table 1.**
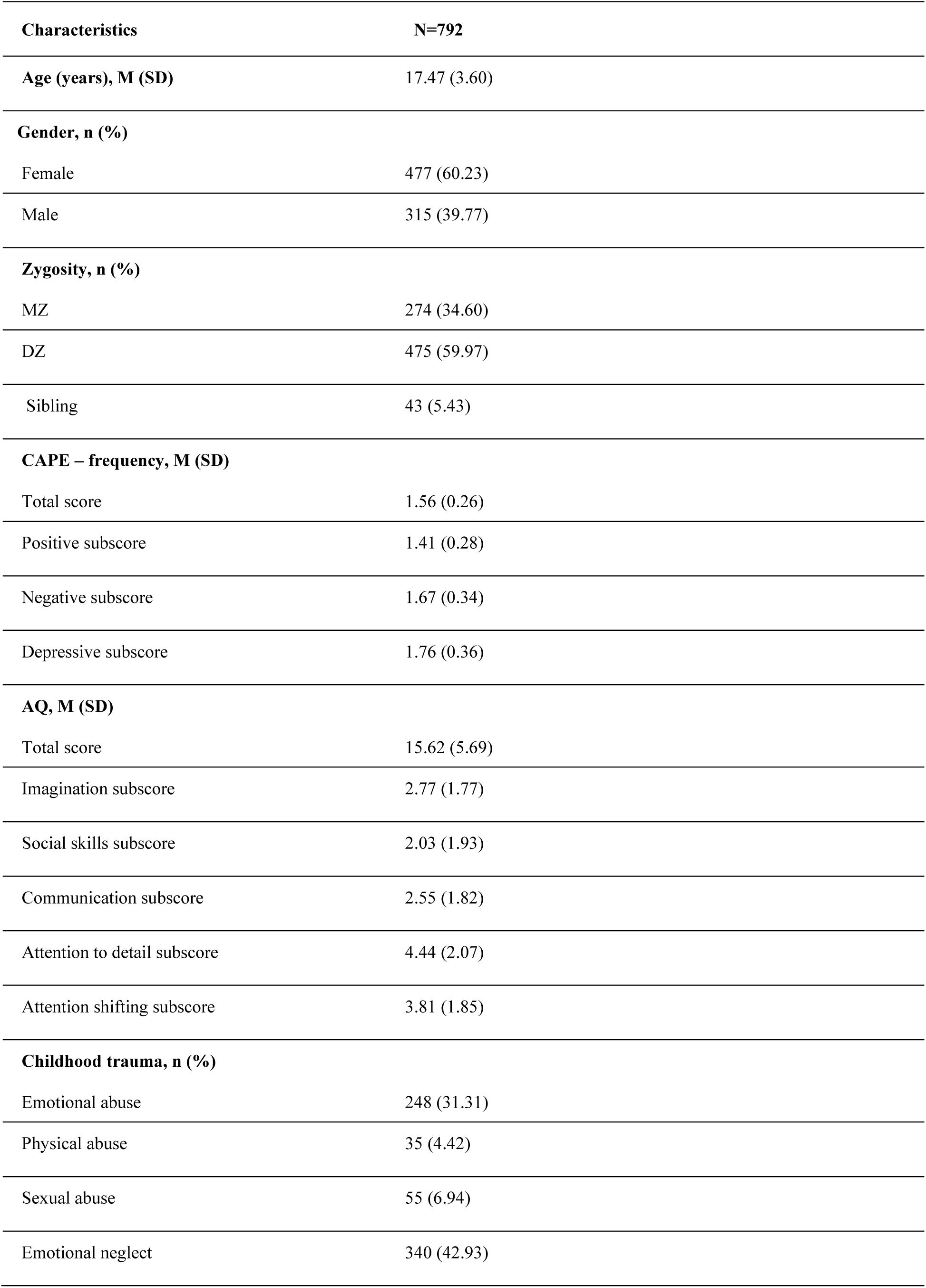
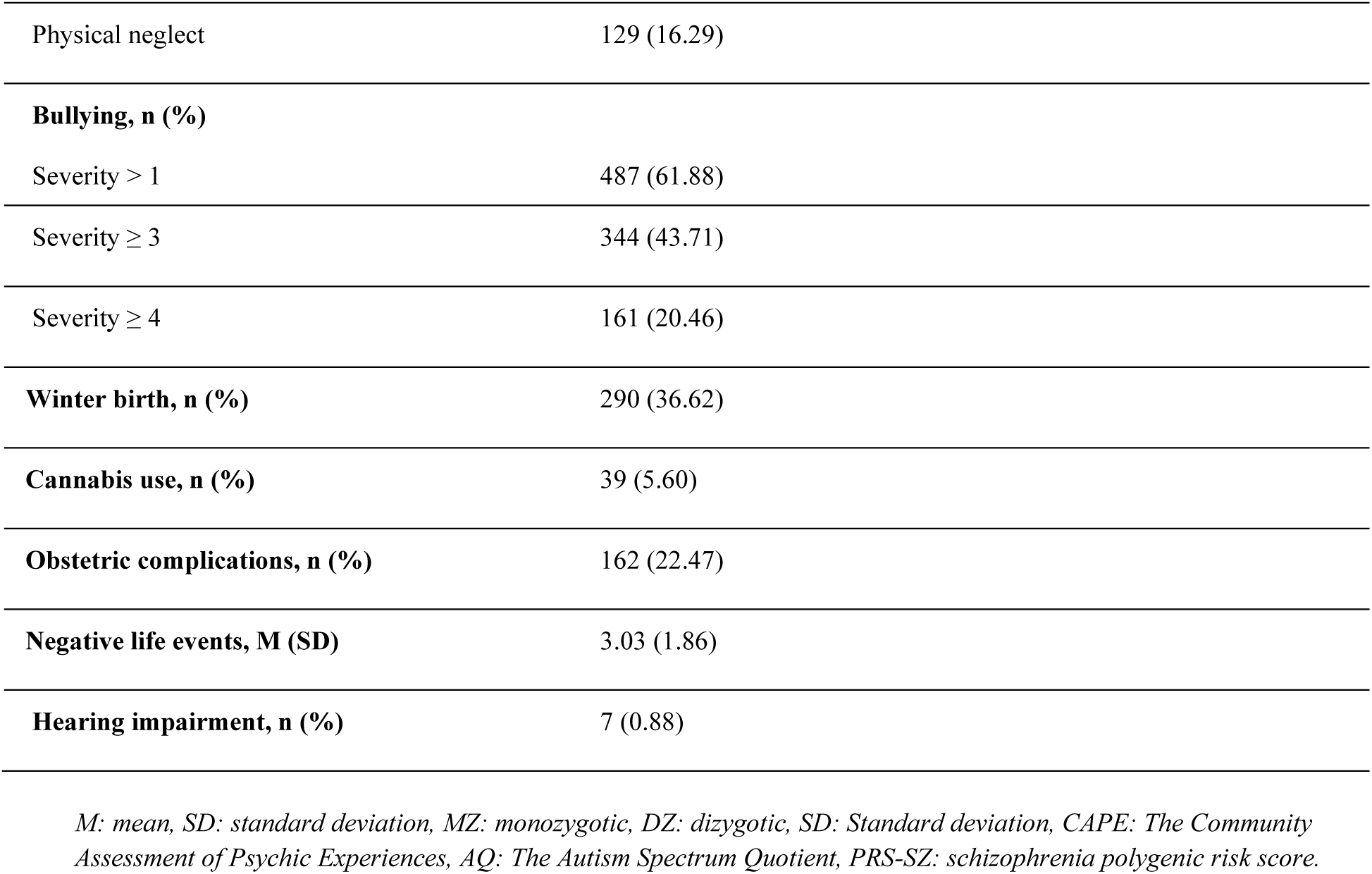
Sample Characteristics (N = 792)

| Characteristics | N=792 |
| --- | --- |
| **Age (years), M (SD)** | 17.47 (3.60) |
| **Gender, n (%)** |  |
| Female | 477 (60.23) |
| Male | 315 (39.77) |
| **Zygoty, n (%)** |  |
| MZ | 274 (34.60) |
| DZ | 475 (59.97) |
| Sibling | 43 (5.43) |
| **CAPE – frequency, M (SD)** |  |
| Total score | 1.56 (0.26) |
| Positive subscore | 1.41 (0.28) |
| Negative subscore | 1.67 (0.34) |
| Depressive subscore | 1.76 (0.36) |
| **AQ, M (SD)** |  |
| Total score | 15.62 (5.69) |
| Imagination subscore | 2.77 (1.77) |
| Social skills subscore | 2.03 (1.93) |
| Communication subscore | 2.55 (1.82) |
| Attention to detail subscore | 4.44 (2.07) |
| Attention shifting subscore | 3.81 (1.85) |
| **Childhood trauma, n (%)** |  |
| Emotional abuse | 248 (31.31) |
| Physical abuse | 35 (4.42) |
| Sexual abuse | 55 (6.94) |
| Emotional neglect | 340 (42.93) |
| Physical neglect | 129 (16.29) |
| **Bullying, n (%)** |  |
| Severity > 1 | 487 (61.88) |
| Severity ≥ 3 | 344 (43.71) |
| Severity ≥ 4 | 161 (20.46) |
| **Winter birth, n (%)** | 290 (36.62) |
| **Cannabis use, n (%)** | 39 (5.60) |
| **Obstetric complications, n (%)** | 162 (22.47) |
| **Negative life events, M (SD)** | 3.03 (1.86) |
| **Hearing impairment, n (%)** | 7 (0.88) |

*M: mean, SD: standard deviation, MZ: monozygotic, DZ: dizygotic, SD: Standard deviation, CAPE: The Community Assessment of Psychic Experiences, AQ: The Autism Spectrum Quotient, PRS-SZ: schizophrenia polygenic risk score.*

### Associations of ATs and psychosis-associated risk factors with PEs

Autistic traits were significantly associated with total CAPE (Table 2, B: 0.12, 95% CI: 0.10 to 0.14, P < 0.001). While PRS-SZ was not associated with total CAPE (B: 0.01, 95% CI: - 0.01 to -0.03, P = 0.33), all five CT subtypes, bullying, and negative life events showed significant positive associations with total CAPE (all P < 0.001). No significant associations between other psychosis risk factors and total CAPE were found (Table 2).

**Table 2.**
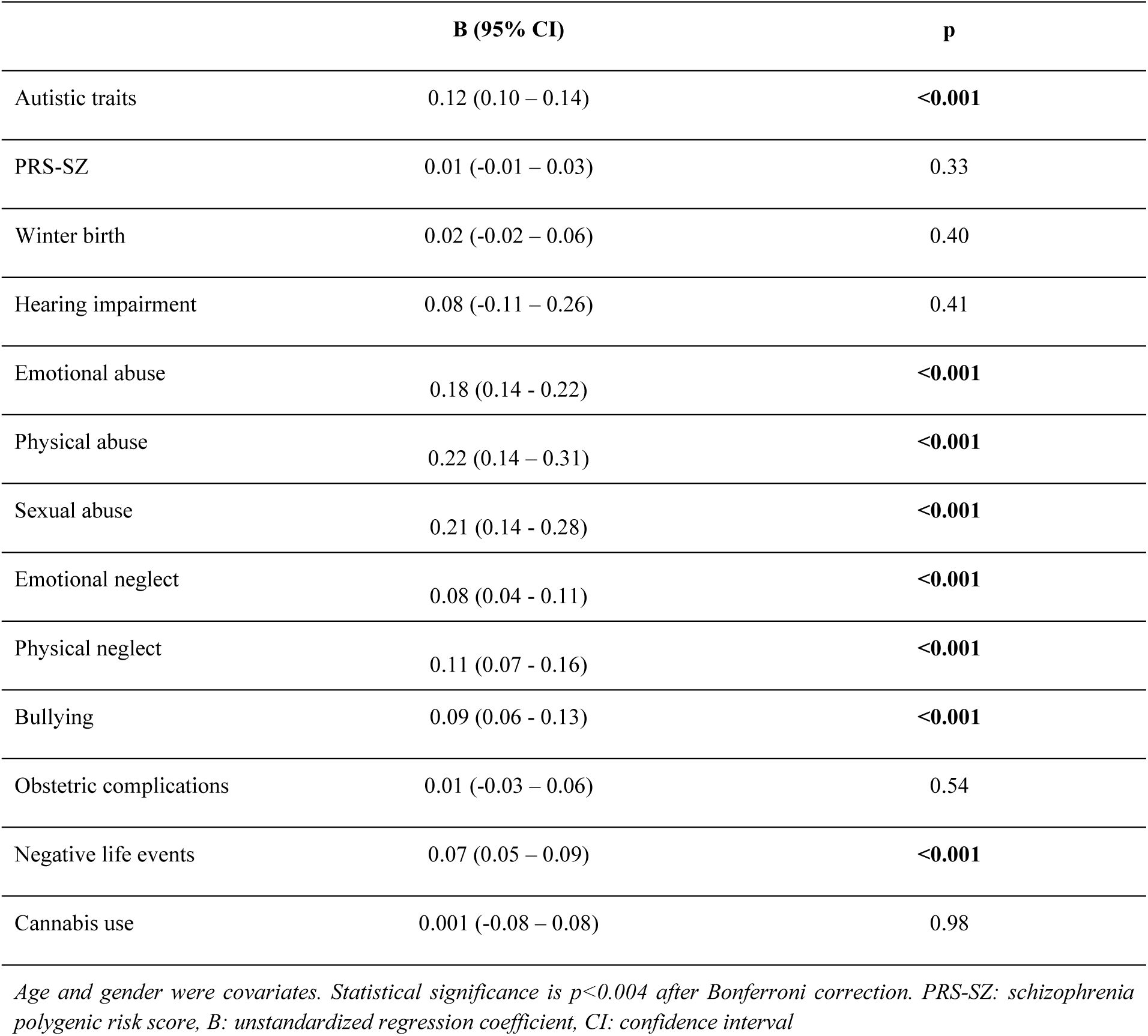
Associations between psychosis-associated risk factors and autistic traits with Pes.

|  | **B (95% CI)** | **p** |
| --- | --- | --- |
| Autistic traits | 0.12 (0.10 – 0.14) | **<0.001** |
| PRS-SZ | 0.01 (-0.01 – 0.03) | 0.33 |
| Winter birth | 0.02 (-0.02 – 0.06) | 0.40 |
| Hearing impairment | 0.08 (-0.11 – 0.26) | 0.41 |
| Emotional abuse | 0.18 (0.14 - 0.22) | **<0.001** |
| Physical abuse | 0.22 (0.14 – 0.31) | **<0.001** |
| Sexual abuse | 0.21 (0.14 - 0.28) | **<0.001** |
| Emotional neglect | 0.08 (0.04 - 0.11) | **<0.001** |
| Physical neglect | 0.11 (0.07 - 0.16) | **<0.001** |
| Bullying | 0.09 (0.06 - 0.13) | **<0.001** |
| Obstetric complications | 0.01 (-0.03 – 0.06) | 0.54 |
| Negative life events | 0.07 (0.05 – 0.09) | **<0.001** |
| Cannabis use | 0.001 (-0.08 – 0.08) | 0.98 |

*Age and gender were covariates. Statistical significance is $p < 0.004$ after Bonferroni correction. PRS-SZ: schizophrenia polygenic risk score, B: unstandardized regression coefficient, CI: confidence interval*

Similar to the main analysis, exploratory analyses of CAPE subscales showed significant positive associations of ATs, all CT subtypes, bullying, and negative life events with all three CAPE subscale scores (Supplementary Tables 3-5).

### Moderations of psychosis-associated risk factors on the associations between ATs and PEs

An interaction analysis revealed that PRS-SZ did not moderate the association between ATs and total CAPE. However, emotional abuse (B: 0.08, 95% CI: 0.05 to 0.11, P < 0.001), physical abuse (B: 0.11, 95% CI: 0.05 to 0.18, P = 0.001), sexual abuse (B: 0.09, 95% CI: 0.03 to 0.15, P = 0.002), and physical neglect (B: 0.06, 95% CI: 0.03 to 0.10, P = 0.001) significantly interacted with ATs in predicting total CAPE. Emotional neglect (B: 0.04, 95% CI: 0.01 to 0.07, P = 0.007) and negative life events (B: 0.007, 95% CI: 0.0005 to 0.014, P = 0.04) significantly interacted with ATs in predicting total CAPE at a nominal level, but the statistical significance was lost after correction for multiple testing (Table 3).

**Table 3.** Interaction effects of psychosis-associated risk factors and ATs on PEs.

|  | **B (95% CI)** | **p** |
| --- | --- | --- |
| PRS-SZ | -0.004 (-0,025 - 0,017) | 0.70 |
| Winter birth | 0.03 (-0.002 – 0.06) | 0.07 |
| Hearing impairment | 0.05 (-0.19 - 0.30) | 0.69 |
| Emotional abuse | 0.08 (0.05 - 0.11) | **<0.001** |
| Physical abuse | 0.11 (0.05 - 0.18) | **0.001** |
| Sexual abuse | 0.09 (0.03 - 0.15) | **0.002** |
| Emotional neglect | 0.04 (0.01 - 0.07) | 0.007 |
| Physical neglect | 0.06 (0.03 - 0.10) | **0.001** |
| Bullying | 0.02 (-0.01 - 0.05) | 0.22 |
| Obstetric complications | -0.001 (-0.040 - 0,038) | 0.94 |
| Negative life events | 0.007 (0.0005 - 0.014) | 0.04 |
| Cannabis use | -0.04 (-0.12 - 0.03) | 0.27 |

*Age and gender were covariates. Statistical significance is $p < 0.004$ after Bonferroni correction. PRS-SZ: schizophrenia polygenic risk score, B: unstandardized regression coefficient, CI: confidence interval*

The marginal plots revealed that the positive association between ATs and total CAPE increased in the presence of CT subtypes (Figure 1). No significant moderations of other psychosis risk factors on the relationship between ATs and total CAPE were found. Exploratory analyses of CAPE subscales showed similar significant interactions between CT subtypes and ATs for CAPE positive subscale (Supplementary Tables 6). However, emotional abuse was the only risk factor significantly interacting with ATs in determining CAPE negative and depressive subscales (Supplementary Tables 7 & 8).

**Figure 1.**
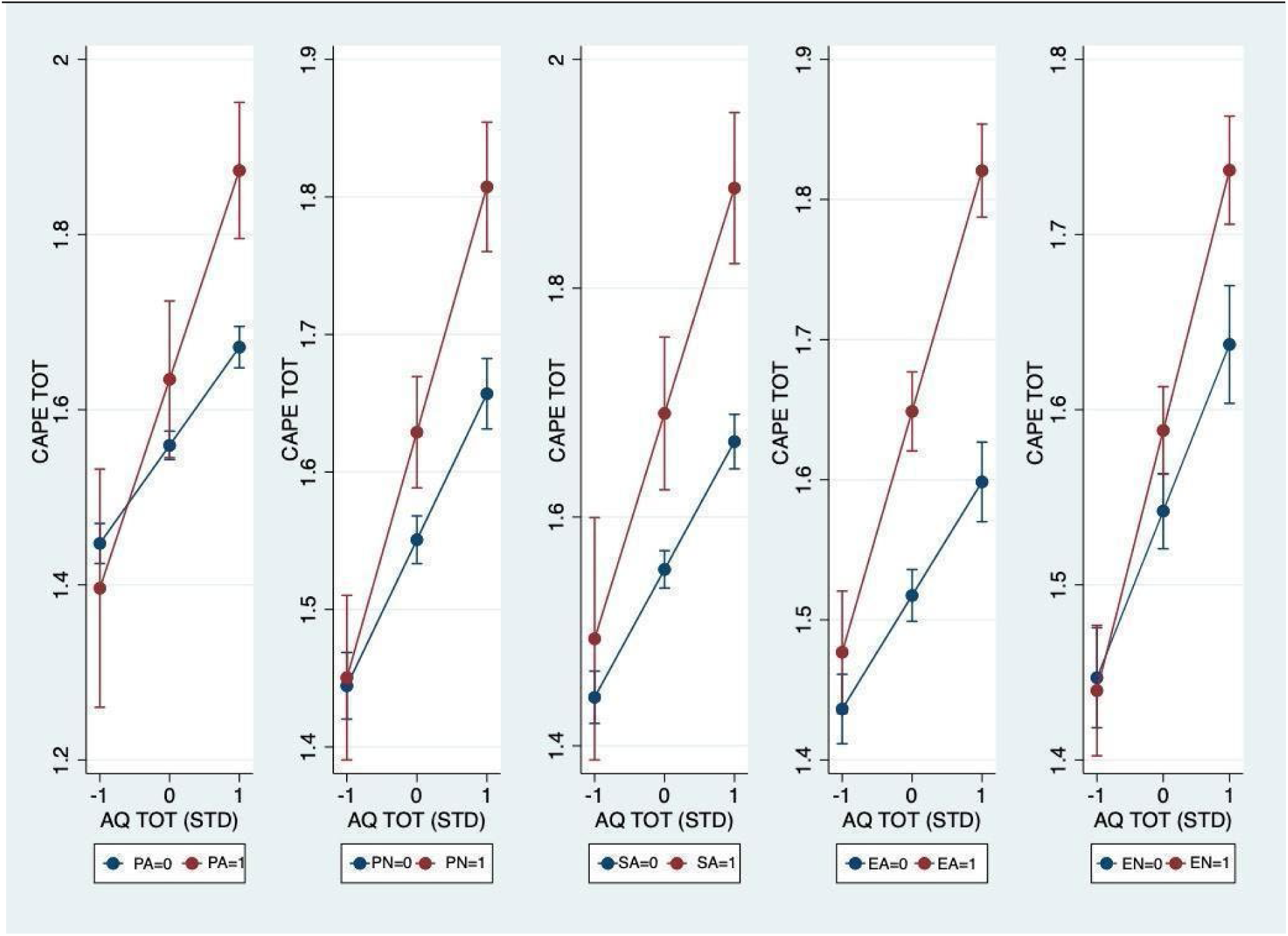
**Interaction effects of childhood trauma subtypes and ATs on Pes** Marginal effect plots based on multilevel linear regression of the interaction between standardized total AQ score (x-axis) and different types of childhood trauma score on total CAPE frequency score(y-axis). CAPE TOT: Total CAPE frequency, AQ TOT (STD): Standardized Total AQ, PA:Physical abuse, PN: Physical neglect, SA:Sexual abuse, EA:Emotional abuse, EN:Emotional neglect

## Discussion

The present study aimed to evaluate the role of psychosis-associated risk factors in moderating the association between ATs and PEs in a general population twin sample. Our primary analyses revealed that ATs significantly interacted with the CT subtypes, particularly emotional abuse, physical abuse, sexual abuse, and physical neglect, on the expression of psychosis, whereas genetic and the other tested environmental risk factors for psychosis did not. Exploratory analyses of psychosis subdomains yielded similar positive interactions between CT subtypes and ATs for positive symptoms, but only emotional abuse significantly interacted with ATs on the expression of depressive and negative symptoms.

We found that CT, but not bullying, significantly moderates the relationship between ATs and PEs. This is partially aligned with the previous research. To date, only few studies have addressed the impact of CT and bullying on the risk of psychosis among individuals with elevated ATs. Two studies have consistently shown that both CT and bullying can mediate the relationship between ATs and PEs^36,37^. Adding to prior evidence, our study suggests that CT, but not bullying, moderates the relationship between ATs and PEs. A possible explanation for this discrepancy between the impact of CT and bullying could be that the perpetrators of CT are usually parents, family members, or relatives^38^, who could have a more profound impact on a child’s life and emotional development than the perpetrators of bullying, who are often peers. External support systems might also be available to help lessen the impact of bullying.

Additionally, bullying tends to be more episodic, whereas chronic and insidious CT may involve prolonged exposure without immediate access to support, possibly leading to a more pervasive and enduring impact. Another speculation could be that CT might contribute to the development of maladaptive coping strategies such as avoidance and rumination^39^, as well as an increased threat anticipation^40^ that could potentially lead to PEs, especially in autistic individuals who already tend to misinterpret threats. While bullying may also have similar effects, its impact may be more prevalent in certain social contexts compared to the broader and more internalized effect of CT.

Our exploratory analyses revealed significant interactions between various CT subtypes and ATs on positive symptoms similar to the overall psychosis expression. However, emotional abuse was the only form of CT that also interacted with ATs in determining negative and depressive symptoms. This is in line with the literature reporting that emotional abuse is a chronic, widespread type of CT and has been found to cause more severe developmental consequences into adulthood, than those caused by other types of CT^41^. Altogether, these findings implied that all forms of CT could amplify the risk of subclinical psychosis, especially positive symptoms in people with high ATs, whereas emotional abuse may exert a broader influence on multiple domains of psychosis expression in this population. Indeed, literature has acknowledged emotional abuse as the form of abuse most frequently linked to mental health problems^42^.

The number of negative life events did not interact with ATs, despite its significant effect on PEs. A recent study found that exposure to family problems and conflicts with peers was associated with increased occurrence of psychotic/manic symptoms in autistic inpatients^43^; however, the relationship between ATs and life events was not studied in non-clinical samples. One possible explanation for this discrepancy is that individuals from our general population sample with high ATs levels, but do not meet the formal criteria for an ASD diagnosis, may cope better with negative life events than those from clinical samples. On the other hand, individuals with higher ATs might already have an elevated baseline of stress perception. Consequently, an increased number of negative life events may not significantly raise their stress levels or lead to PEs, resulting in a less pronounced effect of these events and a weaker association with ATs.

Winter birth, obstetric complications, hearing impairment, and cannabis use were not significantly associated with PEs and did not interact with ATs in predicting PEs. To the best of our knowledge, no studies have ever explored interactions of these factors with ATs on PEs before. First, while some studies suggest a seasonal effect on autism, the findings have been inconsistent, with various studies reporting different seasons of birth, such as fall^44^ or spring births^45,46^ as potential risk factors or no association at all^47,48^. Considering that the previously reported association between winter birth and psychosis expression is generally weak^49,50^, such mixed findings might simply reflect the complex relationship between the season of birth and the risks for psychosis and autism. The level of complexity might require a larger study to detect more subtle effects. Similarly, our limited number of participants with hearing impairment (n=7) and cannabis use (n=39) might offer low statistical power. To this extent, given that ASD and hearing problems frequently co-occur, with a reported ASD prevalence of 9% in children with hearing impairment^51^, further studies on the effect of hearing impairment on psychosis expression in the context of the autistic spectrum are warranted. Also, while existing research on cannabis use in ASD primarily focuses on medical use^52^, the pro-psychotic impact of recreational cannabis exposure among autistic individuals and people with high ATs should not be overlooked, due to disruptive epigenetic effects potentially implicated in pathophysiology of schizophrenia^53,54^.

In contrast to prior reports from larger population-based samples^55,56^, we did not find a significant association between PRS-SZ and PEs, as well as their interaction, which might be attributed to the relatively small sample size of our study. Indeed, other prior studies reporting a null association are also suspected to suffer from inadequate statistical power to detect a potentially weak effect of PRS-SZ on PEs ^57,58^.

### Limitations

Several limitations should be considered. First, childhood adversities, bullying, and life events were evaluated using self-report measures, which may be subjected to recall bias. However, both retrospective and prospective reports of childhood trauma were demonstrated to be linked to mental health issues during adolescence and adulthood^59,60^, supporting the validity of our findings based on retrospective reports. Second, our general population sample allowed us to investigate ATs and PEs as a continuum. However, it limits the generalization to those meeting the full criteria of ASD. Similar analyses in clinical samples should be conducted even though a validated tool for assessing PEs in autistic children remains to be developed^61^. Third, the current study might be prone to type II error for some risk factors with low prevalence or small effect sizes, like hearing impairment, cannabis use, and obstetric complications. Lastly, our study was a cross-sectional analysis; therefore, a temporal relationship between the risk factors and PEs could not be drawn. Further investigation using a longitudinal design is needed for causal inference.

### Conclusions

These findings underscore the significant role of environmental risks in shaping the complex continuum of autism and psychosis. Our findings suggest that identifying individuals with higher ATs and screening them for childhood adversity could facilitate the provision of preventive measures to safeguard them from further environmental risk exposure. These insights not only contribute to our understanding of the intricate links between autism and psychosis but also hold implications for tailoring personalized interventions and treatment plans.

## Data Availability

All data produced in the present study are available upon reasonable request to the authors.

## Funding

The East Flanders Prospective Twin Survey (EFPTS) received support from the Association for Scientific Research in Multiple Births (Belgium) and that the TwinssCan project is funded by the European Community Seventh Framework Program under grant agreement No. HEALTH-F2-2009-241909 (Project EU-GEI). This work was further supported by the Scientific and Technological Research Council of Türkiye (TUBITAK), 2219 International Postdoctoral Research Fellowship under grant number 1059B192302449 to M.K.D., by Ophelia research project, ZonMw under grant number 36340001 to J.V.O. and S.G., the Netherlands Scientific Organisation Vidi award under grant number 91718336 to B.P.F.R., the European Union’s Horizon Europe program, YOUTH-GEMs Project under grant number 01057182 to J.V.O., L.K.P., B.D.L., B.P.F.R., S.G. and A.A.M. and by #NEXTGENERATIONEU (NGEU) from the Ministry of University and Research (MIUR), National Recovery and Resilience Plan (NRRP), project MNESYS (PE0000006) – A Multiscale integrated approach to the study of the nervous system in health and disease to L.F.P. (DN. 1553 11.10.2022). T.P. received research training funding from the Faculty of Medicine Ramathibodi Hospital, Mahidol University. L.K.P. was a recipient of the Kootstra Talent Fellowship of Maastricht University.

## Conflicts of Interest

Marco Colizzi has been a consultant/advisor to GW Pharma Limited, GW Pharma Italy SRL and F. Hoffmann-La Roche Limited, outside of this work.

## Notes

### Author Declarations

The ethics committee research of KU Leuven (Commissie Medische Ethiek van de Universitaire Ziekenhuizen KU Leuven, Nr. B32220107766) gave ethical approval for this work.

