## Supplementary files for "The Moderating Role of Genetic and Environmental Risk Factors for Schizophrenia on the Relationship between Autistic Traits and Psychosis Expression in the General Population"

**Supplementary Figure 1**

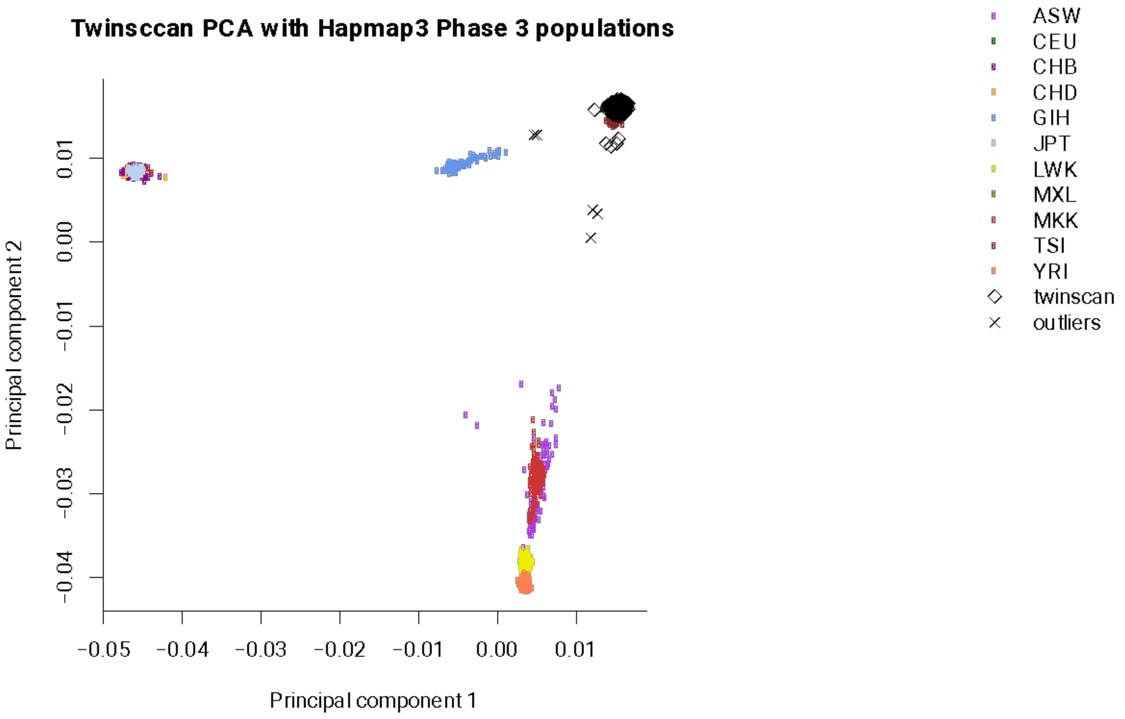


*The first and second principal component of TwinssCan data (with identified ethnic outliers) along with hapmap3 populat*

### Supplementary Tables

**Table-1**. 20 Complex-LD regions and long-range LD regions excluded from PRS analysis.

| Chromosome | Base pair position (start point to end point) |
|---|---|
| 1 | 48000000-52000000 |
| 2 | 86000000-100500000 |
| 2 | 183000000-190000000 |
| 3 | 47500000-50000000 |
| 3 | 83500000-87000000 |
| 5 | 44500000-50500000 |
| 5 | 129000000-132000000 |
| 6 | 25500000-33500000 |
| 6 | 57000000-64000000 |
| 6 | 140000000-142500000 |
| 7 | 55000000-66000000 |
| 8 | 8000000-12000000 |
| 8 | 43000000-50000000 |
| 8 | 112000000-115000000 |
| 10 | 37000000-43000000 |
| 11 | 87500000-90500000 |
| 12 | 33000000-40000000 |
| 20 | 32000000-34500000 |
| 8 | 8135000-12000000 |
| 17 | 40900000-45000000 |

**Table-2**. The number of alleles used for PRS calculation for the TwinssCan data at different P-value thresholds using the different cohorts

| Level | *P*-value threshold | Number of SNPs |
|---|---|---|
| 1 | $5x10^{-8}$ | 168 |
| 2 | $5x10^{-7}$ | 302 |
| 3 | $5x10^{-6}$ | 567 |
| 4 | $5x10^{-5}$ | 1236 |
| 5 | $5x10^{-4}$ | 2900 |
| 6 | $5x10^{-3}$ | 8144 |
| 7 | 0.05 | 26635 |
| 8 | 0.1 | 38852 |
| 9 | 0.2 | 56596 |
| 10 | 0.3 | 70225 |
| 11 | 0.4 | 81110 |
| 12 | 0.5 | 89965 |
| 13 | 1 | 113020 |

*Age and gender were covariates. Statistical significance is p<0.004 after Bonferroni correction. PRS-SZ: schizophrenia polygenic risk score, B: unstandardized regression coefficient, CI: confidence interval*

**Table-3.** Associations between psychosis-associated risk factors and autistic traits with CAPE positive subscores

| | **B (95% CI)** | **p** |
|---|---|---|
| Autistic traits | 0.10 (0.08 - 0.12) | **<0.001** |
| PRS-SZ | 0.01 (-0.01- 0.03) | 0.28 |
| Winter birth | 0.01 (-0.19 - 0.20) | 0.28 |
| Hearing impairment | 0.03 (-0.02- 0.07) | 0.96 |
| Emotional abuse | 0.16 (0.12 - 0.20) | **<0.001** |
| Physical abuse | 0.25 (0.16 - 0.34) | **<0.001** |
| Sexual abuse | 0.23 (0.06 - 0.13) | **<0.001** |
| Emotional neglect | 0.06 (0.06 - 0.13) | **0.004** |
| Physical neglect | 0.12 (0.06 - 0.13) | **<0.001** |
| Bullying | 0.10 (0.06 - 0.14) | **<0.001** |
| Obstetric complications | 0.03 (-0.02 - 0.08) | 0.25 |
| Negative life events | 0.07 (0.05 - 0.08) | **<0.001** |
| Cannabis use | -0.05 (-0.14 - 0.04) | 0.27 |

*Age and gender were covariates. Statistical significance is p<0.004 after Bonferroni correction. PRS-SZ: schizophrenia polygenic risk score, B: unstandardized regression coefficient, CI: confidence interval*

**Table-4.** Associations between psychosis-associated risk factors and autistic traits with CAPE negative subscores

| | B (95% CI) | p |
|---|---|---|
| Autistic traits | 0.16 (0.14 - 0.18) | **<0.001** |
| PRS-SZ | 0.00 (-0.03- 0.03) | 0.99 |
| Winter birth | 0.03 (-0.02- 0.07) | 0.28 |
| Hearing impairment | 0.16 (-0.09 - 0.40) | 0.21 |
| Emotional abuse | 0.18(0.13 - 0.23) | **<0.001** |
| Physical abuse | 0.19(0.08 - 0.31) | **0.001** |
| Sexual abuse | 0.16(0.07 - 0.26) | **<0.001** |
| Emotional neglect | 0.10(0.05 - 0.14) | **<0.001** |
| Physical neglect | 0.12 (0.05 - 0.18) | **<0.001** |
| Bullying | 0.09 (0.04 - 0.14) | **<0.001** |
| Obstetric complications | -0.002 (-0.06- 0.06) | 0.94 |
| Negative life events | 0.07 (0.04 - 0.09) | **<0.001** |
| Cannabis use | 0.03 (-0.08- 0.15) | 0.59 |

*Age and gender were covariates. Statistical significance is p<0.004 after Bonferroni correction. PRS-SZ: schizophrenia polygenic risk score, B: unstandardized regression coefficient, CI: confidence interval*

**Table-5.** Associations between psychosis-associated risk factors and autistic traits with CAPE depressive subscores

| | **B (95% CI)** | **p** |
|---|---|---|
| Autistic traits | 0.11 (0.09 - 0.14) | **<0.001** |
| PRS-SZ | 0.02 (0.00- 0.05) | 0.07 |
| Winter birth | 0.03 (-0.02- 0.09) | 0.25 |
| Hearing impairment | 0.15 (-0.11 - 0.41) | 0.25 |
| Emotional abuse | 0.23 (0.18 - 0.28) | **<0.001** |
| Physical abuse | 0.24 (0.12 - 0.36) | **<0.001** |
| Sexual abuse | 0.26 (0.17 - 0.36) | **<0.001** |
| Emotional neglect | 0.11 (0.06 - 0.16) | **<0.001** |
| Physical neglect | 0.10 (0.03 - 0.17) | **0.004** |
| Bullying | 0.10 (0.05 - 0.15) | **<0.001** |
| Obstetric complications | 0.01 (-0.05- 0.07) | 0.76 |
| Negative life events | 0.10 (0.07 - 0.12) | **<0.001** |
| Cannabis use | 0.09 (-0.03- 0.21) | 0.15 |

*Age and gender were covariates. Statistical significance is p<0.004 after Bonferroni correction. PRS-SZ: schizophrenia polygenic risk score, B: unstandardized regression coefficient, CI: confidence interval*

**Table-6.** Interaction effects of psychosis-associated risk factors and ATs on CAPE positive subscores

| | **B (95% CI)** | **p** |
|---|---|---|
| PRS-SZ | -0.01 (-0.02- 0.01) | 0.25 |
| Winter birth | 0.03 (0.00- 0.07) | 0.08 |
| Hearing impairment | -0.10 (-0.38- 0.18) | 0.48 |
| Emotional abuse | 0.12 (0.08 - 0.15) | **<0.001** |
| Physical abuse | 0.15 (0.07 - 0.22) | **<0.001** |
| Sexual abuse | 0.10 (0.03 - 0.16) | **0.003** |
| Emotional neglect | 0.05 (0.02 - 0.09) | 0.005 |
| Physical neglect | 0.07 (0.03 - 0.11) | **0.002** |
| Bullying | 0.02 (-0.02 - 0.06) | 0.27 |
| Obstetric complications | 0.01 (-0.04 - 0.05) | 0.79 |
| Negative life events | 0.01 (-0.004 - 0.02) | 0.17 |
| Cannabis use | -0.06 (-0.14 - 0.03) | 0.20 |

*Age and gender were covariates. Statistical significance is p<0.004 after Bonferroni correction. PRS-SZ: schizophrenia polygenic risk score, B: unstandardized regression coefficient, CI: confidence interval*

**Table-7.** Interaction effects of psychosis-associated risk factors and ATs on CAPE negative subscores

| | B (95% CI) | p |
|---|---|---|
| PRS-SZ | -0.01 (-0.03 - 0.01) | 0.39 |
| Winter birth | 0.02 (-0.02 - 0.07) | 0.35 |
| Hearing impairment | 0.21 (-0.11 - 0.54) | 0.20 |
| Emotional abuse | 0.09 (0.04 - 0.13) | **<0.001** |
| Physical abuse | 0.08 (-0.01 - 0.17) | 0.08 |
| Sexual abuse | 0.10 (0.02 - 0.18) | 0.01 |
| Emotional neglect | 0.03 (-0.01 - 0.07) | 0.15 |
| Physical neglect | 0.06 (0.01 - 0.11) | 0.03 |
| Bullying | 0.03 (-0.02 - 0.07) | 0.21 |
| Obstetric complications | -0.02 (-0.07 - 0.04) | 0.45 |
| Negative life events | 0.02 (0.003 - 0.04) | 0.02 |
| Cannabis use | -0.03 (-0.13 - 0.08) | 0.60 |

*Age and gender were covariates. Statistical significance is $p<0.004$ after Bonferroni correction. PRS-SZ: schizophrenia polygenic risk score, B: unstandardized regression coefficient, CI: confidence interval*

**Table-8.** Interaction effects of psychosis-associated risk factors and ATs on CAPE depressive subscores

| | B (95% CI) | p |
|---|---|---|
| PRS-SZ | -0.01 (-0.03 - 0.01) | 0.37 |
| Winter birth | 0.04 (-0.01 - 0.09) | 0.09 |
| Hearing impairment | 0.21 (-0.15 - 0.58) | 0.26 |
| Emotional abuse | 0.09 (0.04- 0.14) | **<0.001** |
| Physical abuse | 0.11 (0.01- 0.21) | 0.04 |
| Sexual abuse | 0.05 (-0.03 - 0.14) | 0.22 |
| Emotional neglect | 0.05 (0.01 - 0.10) | 0.02 |
| Physical neglect | 0.08 (0.02 - 0.13) | 0.009 |
| Bullying | 0.01 (-0.04 - 0.06) | 0.58 |
| Obstetric complications | 0.02 (-0.04 - 0.07) | 0.56 |
| Negative life events | 0.01 (-0.004 - 0.03) | 0.12 |
| Cannabis use | -0.05 (-0.17 - 0.06) | 0.38 |

*Age and gender were covariates. Statistical significance is p<0.004 after Bonferroni correction. PRS-SZ: schizophrenia polygenic risk score, B: unstandardized regression coefficient, CI: confidence interval*
